# Changing probability of experiencing food insecurity by socioeconomic and demographic groups during the COVID-19 pandemic in the UK

**DOI:** 10.1101/2020.11.10.20229278

**Authors:** Jonathan Koltai, Veronica Toffolutti, Martin Mckee, David Stuckler

## Abstract

**Background:** Food supply concerns have featured prominently in the UK response to the COVID-19 pandemic. We assess changes in food insecurity in the UK population from April to July 2020.

**Method:** We analyze 11,095 respondents from the April through July waves of the Understanding Society COVID-19 longitudinal study survey linked with Wave 9 of the UK Understanding Society study. Food insecurity was defined as having used a food bank in the last 4 weeks; being hungry but not eating in the last week; or not able to eat healthy and nutritious food in the last week. Unadjusted estimates to examine changes in population prevalence and logistic regression were used to assess the association between employment transitions and food insecurity.

**Findings:** The prevalence of reporting at least one form of food insecurity rose from 7·1% in April to 20·2% by July 2020. Some of the largest increases were among Asian respondents (22·91 percentage points), the self-employed (15·90 percentage points), and 35-44-year-olds (17·08 percentage points). In logistic regression models, those moving from employment to unemployment had higher odds of reporting food insecurity relative to furloughed individuals (OR = 2·23; 95% CI: 1·20–4·131) and to the persistently employed (OR=2·38; 95% CI: 1·33–4·27), adjusting for sociodemographic characteristics. Furloughed individuals did not differ significantly in their probability of experiencing food insecurity compared to the persistently employed (OR=1·07; 95% CI: 0·83 to 1·37).

**Interpretation:** Food insecurity has increased substantially in the UK. Steps are needed to provide subsidies or food support to vulnerable groups.

Evidence before this study
We searched Google Scholar with the terms “COVID-19” and “food insecurity” and “UK”; and “food insecurity” and “UK” and “coronavirus”, published between January 1^st^ and October 31^st^, 2020. One cross-sectional report was identified, which found higher levels of food insecurity in early April 2020 relative to 2018. Importantly, the report relied on items used to measure food insecurity that referred to a 12-month time span in 2018 and then a 30-day time span in April 2020, a potential source of bias for examining changes in population prevalence over time.

Added value of this study
Here we provide the first longitudinal national probability study that tracks temporal changes in population prevalence of food insecurity several months following the initial COVID-19-related lockdown measures in the UK. The prevalence of food insecurity rose for all socioeconomic and demographic and groups from April to July 2020, but did so for some more than others. Some of the largest increases in food insecurity were among Asian respondents, the self-employed, respondents aged 35-44, and those living in Scotland, London, and the North West of England. At the individual level, losing employment was associated with a higher odds of food insecurity compared to those furloughed under the Coronavirus Job Retention Scheme and the persistently employed. Importantly, furloughed individuals did not differ in their probability of food insecurity relative to the persistently employed.

Implications of all the available evidence
This study documents an alarming increase in food insecurity in the United Kingdom during the pandemic, with important implications for policy. While Coronavirus the Job Retention Scheme appeared to have conferred some protection, it is clear that not enough has been done to mitigate overall increases food insecurity in the UK. Steps are needed to provide subsidies or food support, especially since during the pandemic emergency food assistance may not be readily accessible. Taken together our results show that, while COVID is first of all a health crisis, it also has potential to become an escalating social and economic crisis if steps are not taken to protect the weak.

## Introduction

Concerns about food supply have featured prominently in the United Kingdom’s response to the COVID-19 pandemic.^1^ In March 2020, the media reported fights breaking out as people attempted to stock up on rapidly diminishing supplies.^2^ Supermarket shelves emptied and food producers asked how they could continue to supply them without placing their workers at risk. The immediate panic resolved but soon gave way to concerns about the many families who, until then, had been just about coping. Austerity measures adopted since 2010 had left many living a precarious existence characterised by insecure employment, income, and in some cases food and shelter.^3^ Many were dependent on the growing number of foodbanks^4^ and, for those who qualified, free school meals for their children. As schools closed, many families found that they had to find additional food for their children, even though many were facing loss of income or employment. In 2019/20 over 1.4 million school children in England, around 15% of the total, received free school meals.^5^ While the rules of entitlement vary among the four nations of the United Kingdom, those whose families are receiving certain benefits are likely to be eligible. One recent study estimated that half of free school meal eligible children could not access the scheme in April 2020.^6^

Food insecurity has become highly politically contentious in the UK. When schools closed in March 2020, a prominent football player, Marcus Rashford, whose family had depended on foodbanks when he was a child, established a charity to deliver food packages to children who were no longer getting school meals. The four governments in the United Kingdom subsequently established schemes to support these children, with varying degrees of success, but the United Kingdom government, which is responsible for education policy in England, rejected continuing its scheme over the summer. In June, Rashford wrote an open letter to the government^7^ and, a day later, the government reversed its decision. However, as of October, England declined to continue its scheme during school holidays, unlike in the other three countries. An opposition Bill calling for it to be reinstated was defeated, although this provoked a split in the Conservative parliamentary party, with some Members of Parliament voting with the opposition^8^ while others made a series of widely criticized comments blaming parents and suggesting that some food parcels were being diverted to pay for illegal drugs.^9^ Over 2,000 British paediatricians wrote an open letter calling on the UK government to match the policies in the other three nations^10^ and a petition to “End child food poverty”, set up by Rashford, has attracted almost 1 million signatures.^11^ Yet while the debate about food insecurity has become extremely acrimonious, it has not always been underpinned by data and most of the times the data used belong to very selective groups such as food bank organizations.

We report the findings of a study that seeks to inform this debate, asking about how food insecurity in the UK has changed during the pandemic, and how this has impacted on different groups within society.

## Methods

Our objective was to assess whether the prevalence of food insecurity has risen in the UK from April to July 2020, for which groups, and to assess whether employment transitions (Employed to furloughed, or employed to unemployed) were associated with higher food insecurity at the individual level.

### Source of data

Data for this study are derived from the UK Understanding Society study (or UK Household Longitudinal Study). The Understanding Society study is a panel survey of more than 40,000 households beginning in 2009, based on a clustered stratified probability sample of UK households, described in detail in previous research.^12 13^

Following the onset of the pandemic, the Understanding Society COVID-19 study was collected via online survey between 24th and 30th April 2020.^14^ The response rate for this wave was 41·2% of those who took part in Waves 8 or 9. When considering only Wave 9 participants, interviews were completed (full and partial) by 15,835 of 32,596 Wave 9 respondents, representing a response rate of 48·6%. A total of 13,754 individuals then participated in the July web survey. The survey weights in the Understanding Society COVID-19 study extend the weighting strategy used in the Understanding Society annual survey, which adjusts for unequal selection probabilities as well as differential nonresponse.

For prevalence analyses, we used data from the April and July waves of the Understanding Society COVID-19 prospective cohort study, linked with socioeconomic and demographic characteristics from Wave 9 (2017-19) of the Understanding Society study (flowcharts in the appendix describe sample generation). This yielded an analytic sample of n = 11,095 individuals for analyses of shifts in the prevalence of food insecurity from April to July 2020.

The outcome of interest was food insecurity, defined as meeting one of the following criteria: having used a food bank in the last 4 weeks; being hungry but not eating in the last week; or not able to eat healthy and nutritious food in the last week. This was converted to a dichotomous variable, which was yes if the respondent answered in the affirmative to any of these items and no otherwise. These items were adapted from the UN Global Food Insecurity Experience Scale.

### Statistical modelling

To quantify changes in the probability of food insecurity, we report unadjusted estimates in Table 1, as well as unadjusted versus adjusted estimates in Figures 1-3, controlling for several potential socio-demographic confounders: age, highest qualification in 2017-19, total net equivalized household income in 2017-19, gender, race/ethnicity, number of school-aged children at home, cohabitation status, government office region, employment status. All estimates adjust for sampling weights for representativeness to the UK population, complex sampling design, and non-random attrition.

**Table 1:**
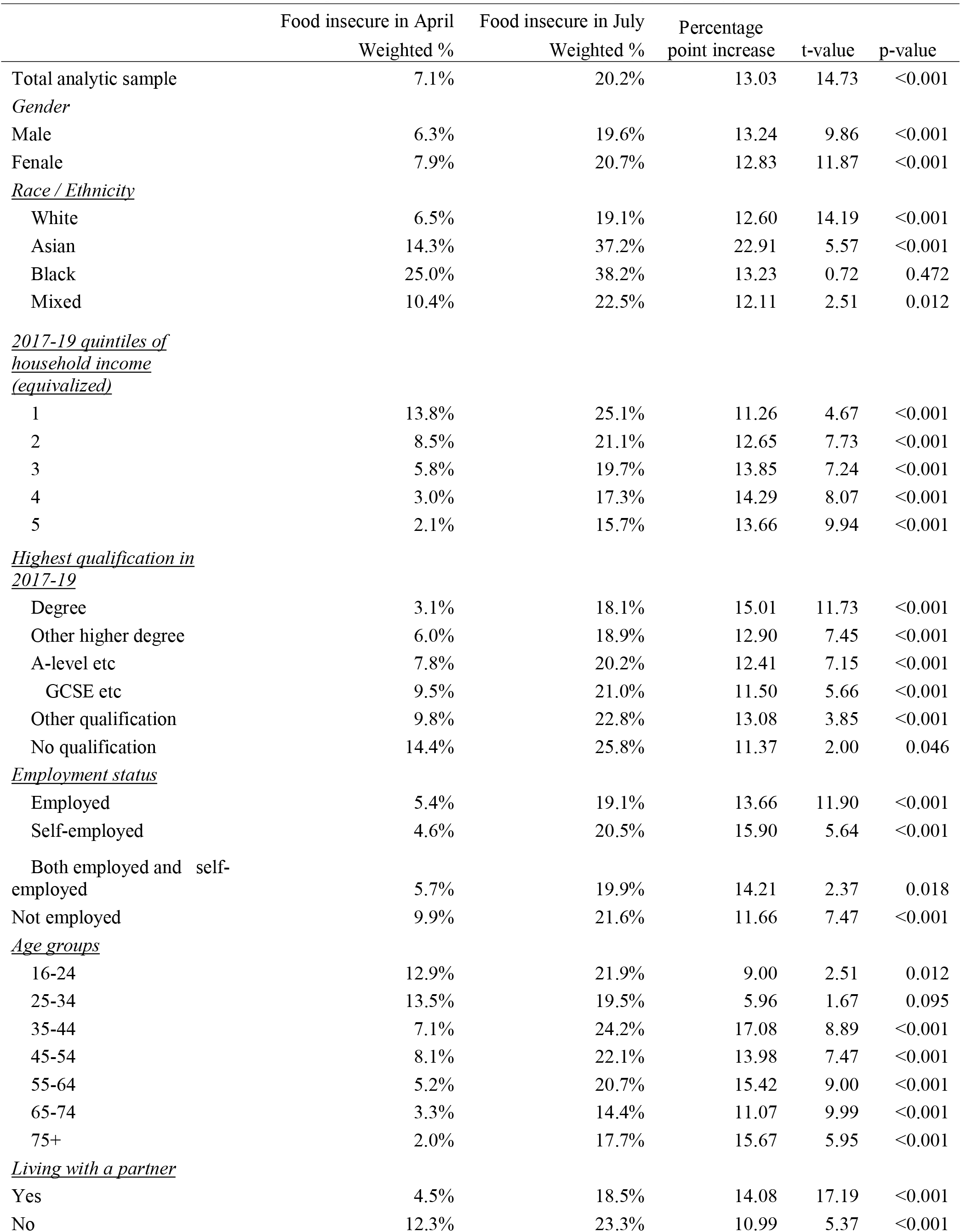
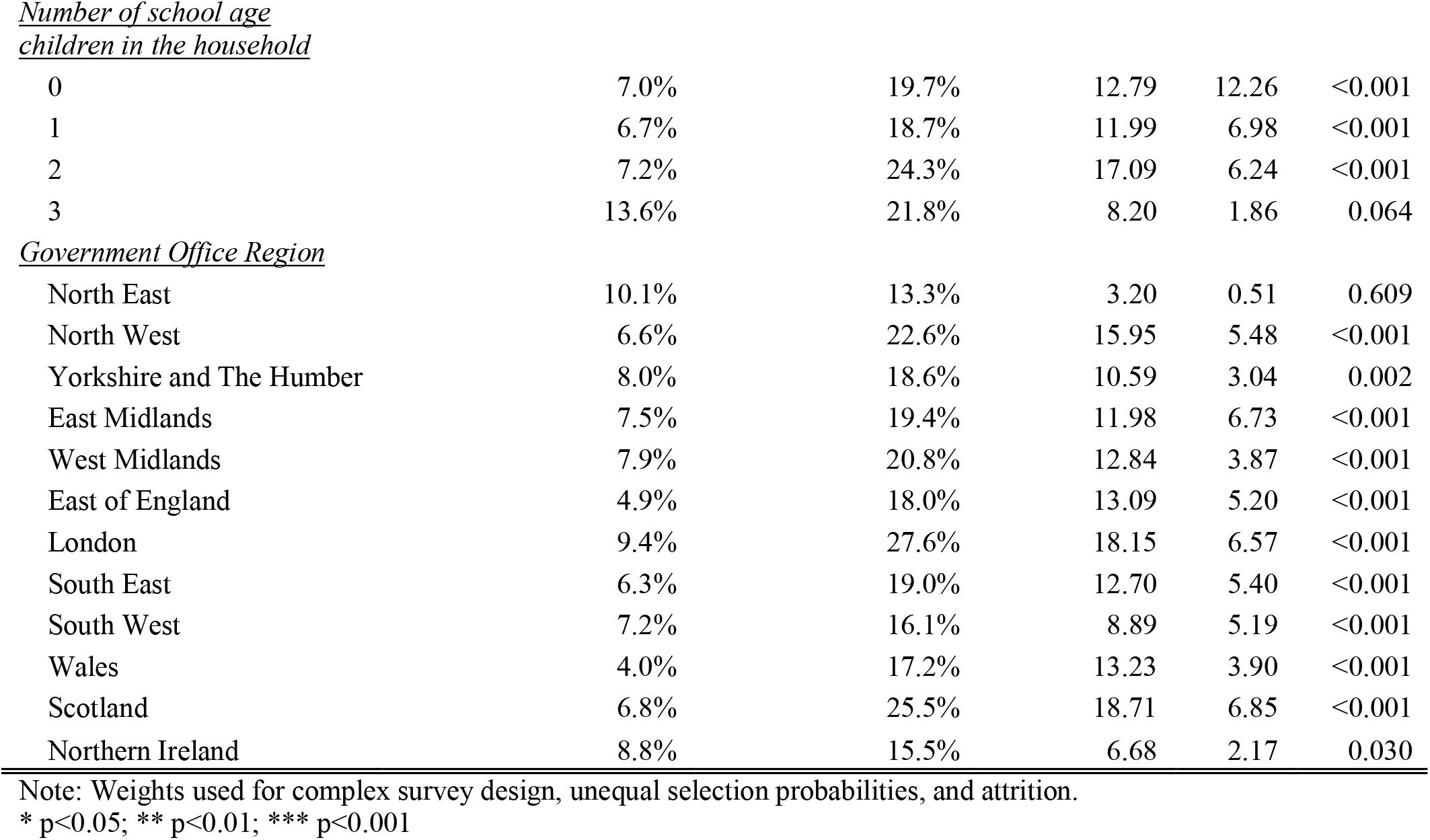
Prevalence of food insecurity across socioeconomic and demographic groups (N=11,095)

|  | Food insecure in April<br>Weighted % | Food insecure in July<br>Weighted % | Percentage<br>point increase | t-value | p-value |
| --- | --- | --- | --- | --- | --- |
| Total analytic sample | 7.1% | 20.2% | 13.03 | 14.73 | <0.001 |
| *Gender* |  |  |  |  |  |
| Male | 6.3% | 19.6% | 13.24 | 9.86 | <0.001 |
| Female | 7.9% | 20.7% | 12.83 | 11.87 | <0.001 |
| *Race / Ethnicity* |  |  |  |  |  |
| White | 6.5% | 19.1% | 12.60 | 14.19 | <0.001 |
| Asian | 14.3% | 37.2% | 22.91 | 5.57 | <0.001 |
| Black | 25.0% | 38.2% | 13.23 | 0.72 | 0.472 |
| Mixed | 10.4% | 22.5% | 12.11 | 2.51 | 0.012 |
| *2017-19 quintiles of*<br>*household income*<br>*(equivalized)* |  |  |  |  |  |
| 1 | 13.8% | 25.1% | 11.26 | 4.67 | <0.001 |
| 2 | 8.5% | 21.1% | 12.65 | 7.73 | <0.001 |
| 3 | 5.8% | 19.7% | 13.85 | 7.24 | <0.001 |
| 4 | 3.0% | 17.3% | 14.29 | 8.07 | <0.001 |
| 5 | 2.1% | 15.7% | 13.66 | 9.94 | <0.001 |
| *Highest qualification in*<br>*2017-19* |  |  |  |  |  |
| Degree | 3.1% | 18.1% | 15.01 | 11.73 | <0.001 |
| Other higher degree | 6.0% | 18.9% | 12.90 | 7.45 | <0.001 |
| A-level etc | 7.8% | 20.2% | 12.41 | 7.15 | <0.001 |
| GCSE etc | 9.5% | 21.0% | 11.50 | 5.66 | <0.001 |
| Other qualification | 9.8% | 22.8% | 13.08 | 3.85 | <0.001 |
| No qualification | 14.4% | 25.8% | 11.37 | 2.00 | 0.046 |
| *Employment status* |  |  |  |  |  |
| Employed | 5.4% | 19.1% | 13.66 | 11.90 | <0.001 |
| Self-employed | 4.6% | 20.5% | 15.90 | 5.64 | <0.001 |
| Both employed and self-employed | 5.7% | 19.9% | 14.21 | 2.37 | 0.018 |
| Not employed | 9.9% | 21.6% | 11.66 | 7.47 | <0.001 |
| *Age groups* |  |  |  |  |  |
| 16-24 | 12.9% | 21.9% | 9.00 | 2.51 | 0.012 |
| 25-34 | 13.5% | 19.5% | 5.96 | 1.67 | 0.095 |
| 35-44 | 7.1% | 24.2% | 17.08 | 8.89 | <0.001 |
| 45-54 | 8.1% | 22.1% | 13.98 | 7.47 | <0.001 |
| 55-64 | 5.2% | 20.7% | 15.42 | 9.00 | <0.001 |
| 65-74 | 3.3% | 14.4% | 11.07 | 9.99 | <0.001 |
| 75+ | 2.0% | 17.7% | 15.67 | 5.95 | <0.001 |
| *Living with a partner* |  |  |  |  |  |
| Yes | 4.5% | 18.5% | 14.08 | 17.19 | <0.001 |
| No | 12.3% | 23.3% | 10.99 | 5.37 | <0.001 |
| 0 | 7.0% | 19.7% | 12.79 | 12.26 | <0.001 |
| 1 | 6.7% | 18.7% | 11.99 | 6.98 | <0.001 |
| 2 | 7.2% | 24.3% | 17.09 | 6.24 | <0.001 |
| 3 | 13.6% | 21.8% | 8.20 | 1.86 | 0.064 |
| Government Office Region |  |  |  |  |  |
| North East | 10.1% | 13.3% | 3.20 | 0.51 | 0.609 |
| North West | 6.6% | 22.6% | 15.95 | 5.48 | <0.001 |
| Yorkshire and The Humber | 8.0% | 18.6% | 10.59 | 3.04 | 0.002 |
| East Midlands | 7.5% | 19.4% | 11.98 | 6.73 | <0.001 |
| West Midlands | 7.9% | 20.8% | 12.84 | 3.87 | <0.001 |
| East of England | 4.9% | 18.0% | 13.09 | 5.20 | <0.001 |
| London | 9.4% | 27.6% | 18.15 | 6.57 | <0.001 |
| South East | 6.3% | 19.0% | 12.70 | 5.40 | <0.001 |
| South West | 7.2% | 16.1% | 8.89 | 5.19 | <0.001 |
| Wales | 4.0% | 17.2% | 13.23 | 3.90 | <0.001 |
| Scotland | 6.8% | 25.5% | 18.71 | 6.85 | <0.001 |
| Northern Ireland | 8.8% | 15.5% | 6.68 | 2.17 | 0.030 |

Note: Weights used for complex survey design, unequal selection probabilities, and attrition.
\* p<0.05; \*\* p<0.01; \*\*\* p<0.001

**Figure 1.**
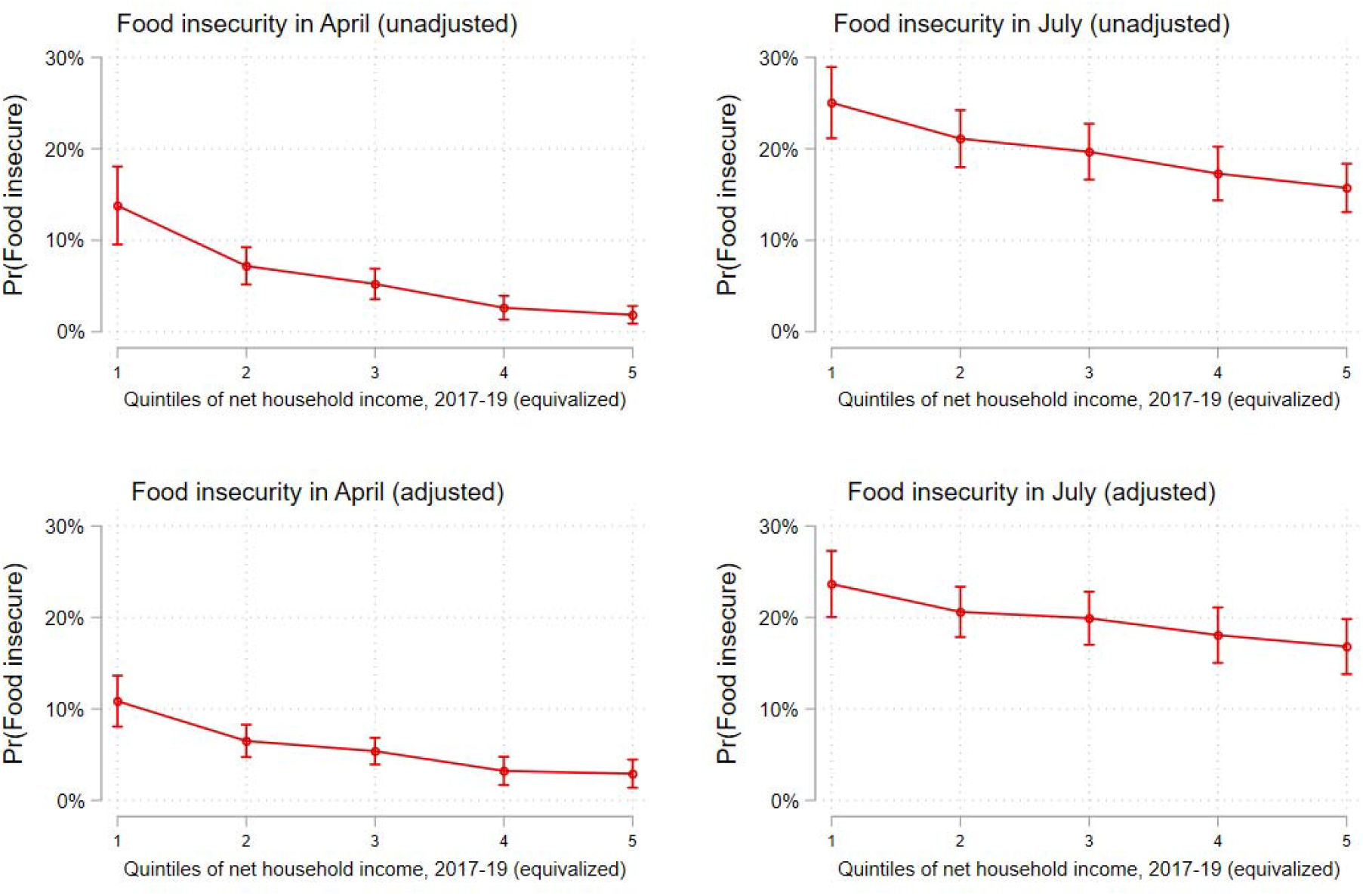
Shift in the probability of experiencing food insecurity across quintiles of household incomes in 2017-19 (equivalized), April-July 2020 Note: Estimates are marginal effects derived from a logistic regression model that controls for age, gender, race/ethnicity, number of children at home, highest qualification in 2017-19, and geographic region. Complex sampling design weights that adjust for non-random attrition are taken into account.

**Figure 2.**
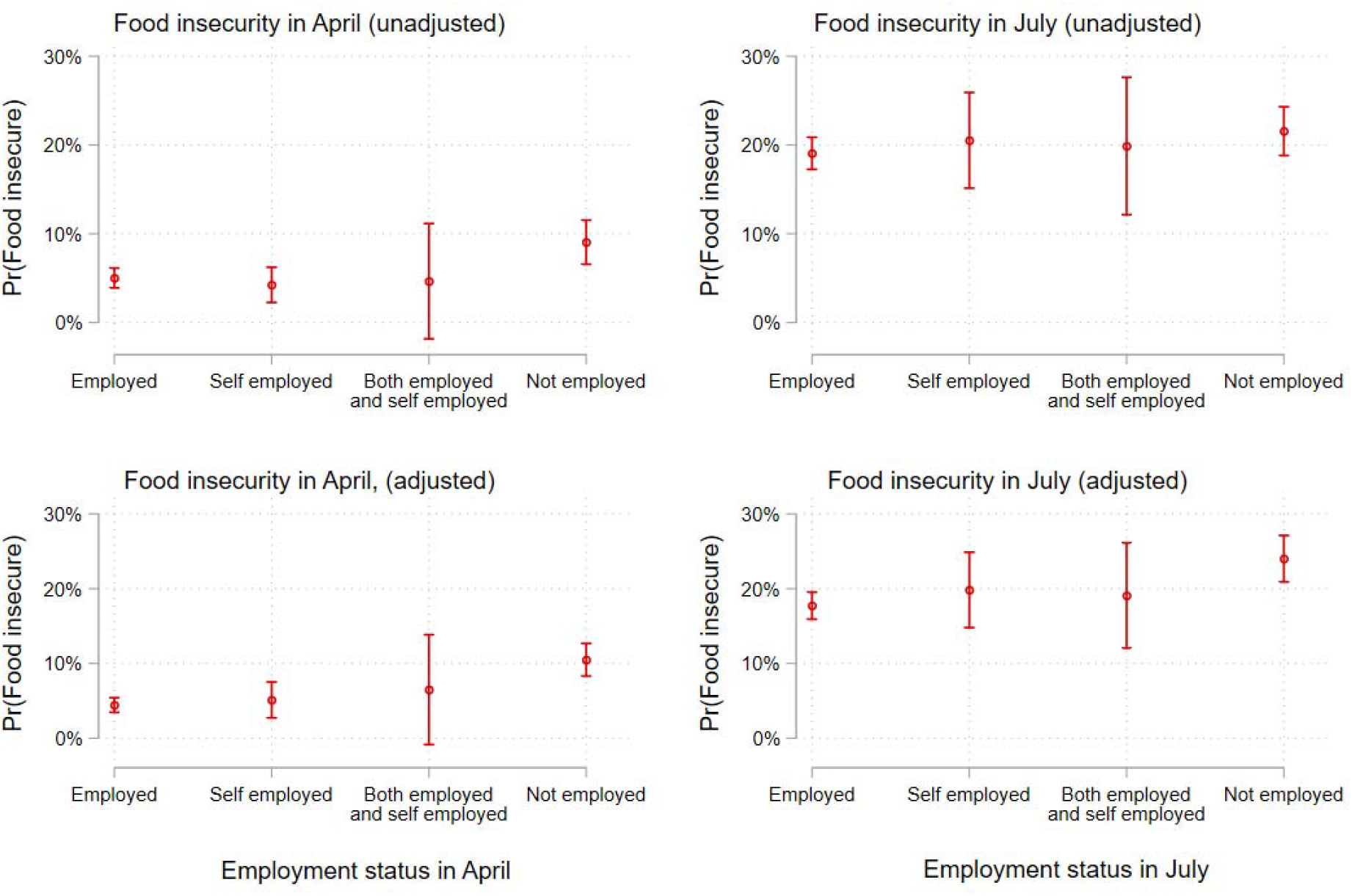
Shift in the probability of experiencing food insecurity by employment status, April-July 2020 Note: Estimates are marginal effects derived from a logistic regression model that controls for age, gender, race/ethnicity, number of children at home, highest qualification in 2017-19, equivalized household income in 2015-17, and geographic region. Complex sampling design weights that adjust for non-random attrition are taken into account.

**Figure 3.**
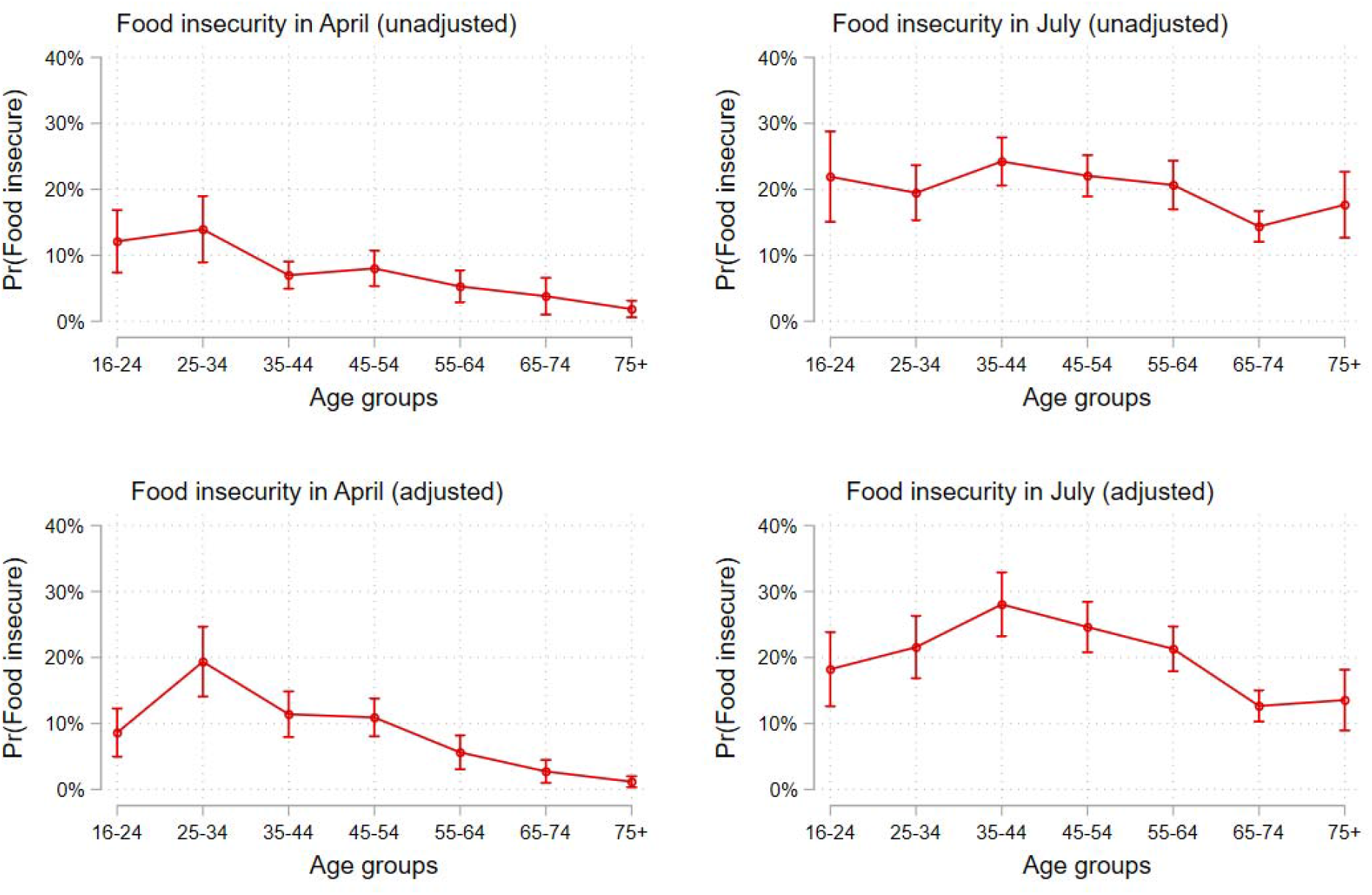
Shift in the probability of experiencing food insecurity by age groups, April-July 2020 Note: Estimates are marginal effects derived from a logistic regression model that controls for gender, race/ethnicity, number of children at home, highest qualification in 2017-19, equivalized household income in 2015-17, and geographic region. Complex sampling design weights that adjust for non-random attrition are taken into account.

For the second objective, we incorporate the May and July waves of the Understanding Society COVID-19 study to identify a series of employment transitions among those who were employed at baseline (as assessed in a retrospective question relating to January / February) including those who were employees, self-employed, or both (number employed at baseline = 5,647). There were three groups. The first were in employment throughout the period January/February to July (n = 3,881). The second were furloughed, which at that time meant that they stopped working but remained in employment while being paid 80% of their previous income, up to a limit (up to a maximum salary of £2,500 per month for employees up to £7,500 for self-employed), and those eligible for the equivalent self-employment support scheme. These were employed in January/February, then furloughed under the Coronavirus Job Retention Scheme or eligible for the self-employment support scheme in either April, May, June, OR July (N = 1,497). The third group transitioned from employed to out of employment. They were employed in January/February, then no longer employed in either April, May, June, OR July (N = 269).

The analysis began with a description of patterns of food insecurity by the main sociodemographic covariates followed by logistic regression modelling, incorporating weights used for complex survey design, unequal selection probabilities, and attrition.

## Results

Unadjusted prevalence estimates for changes in the prevalence of food insecurity between April and July are described in Table 1. There has been a marked rise in food insecurity during the pandemic. In April 2020, 7·1% of respondents reported at least one form of food insecurity. By July, this had increased to 20·2%, a 13·03-percentage point increase (t-value=14·73; p < 0·001). The experience of food insecurity varied substantially within the population. Some of the largest increases in food insecurity were seem among Asian respondents (22·91 percentage points; t-value 5·57; p < 0·001), the self-employed (15·90 percentage points t-value 5·64; p < 0·001), respondents aged 35-44 (17·08 percentage points; t-value 8·89; p < 0·001), and those living in Scotland, London, and the North West of England and Midlands (18·71 percentage points, 18·15 percentage points, and 15·95 percentage points, 11·98 percentage points for East Midlands and 12·84 percentage points for West Midlands respectively, all t-values significant at p < 0·001).

Figures 1-3 illustrate shifts in the prevalence of food insecurity across key socio-demographic groups, presenting both unadjusted and adjusted estimates. In April, food insecurity was, as expected, higher among those in the lowest quintile of household income (Figure 1), although the gradient diminished in the fully adjusted model and no statistically significant differences can be found among the highest four quintiles. By July, the probability of experiencing food insecurity had increased by more than twofold for all quintiles, with gradients in the adjusted and unadjusted model similar to those in April. Looked at by employment status, again, as expected, insecurity was greatest among the unemployed, and very low for those in any form of employment (Figure 2). Again, by July, this had increased markedly for all groups, but especially for the unemployed.

When looked at by age, the picture was more complex (Figure 3). In the adjusted model, it was highest among those aged 25 to 34 in April, but among those aged 35 to 44 in July. The increase was also marked among those aged 55-64 (15·42 percentage points; t-value = 9·00; p < 0·001), 65-74 (11·07 percentage points increase; t-value=9·99; p<0·001) and those aged 75 and older (15·67 percentage points; t-value = 5·95; p <0·001).

Turning to those experiencing employment transitions (Figure 4), those moving from employment to unemployment had higher odds of reporting food insecurity relative to furloughed individuals (OR = 2·23; p < 0·05; 95% CI: 1·20 to 4·131) and to the persistently employed (OR = 2·38; p < 0·01; 95% CI: 1·33 to 4·27), adjusting for age, highest qualification in 2017-19, net household income in 2017-19 (equivalized), gender, race/ethnicity, number of school-aged children at home, cohabitation status, government office region, and employment status.

**Figure 4.**
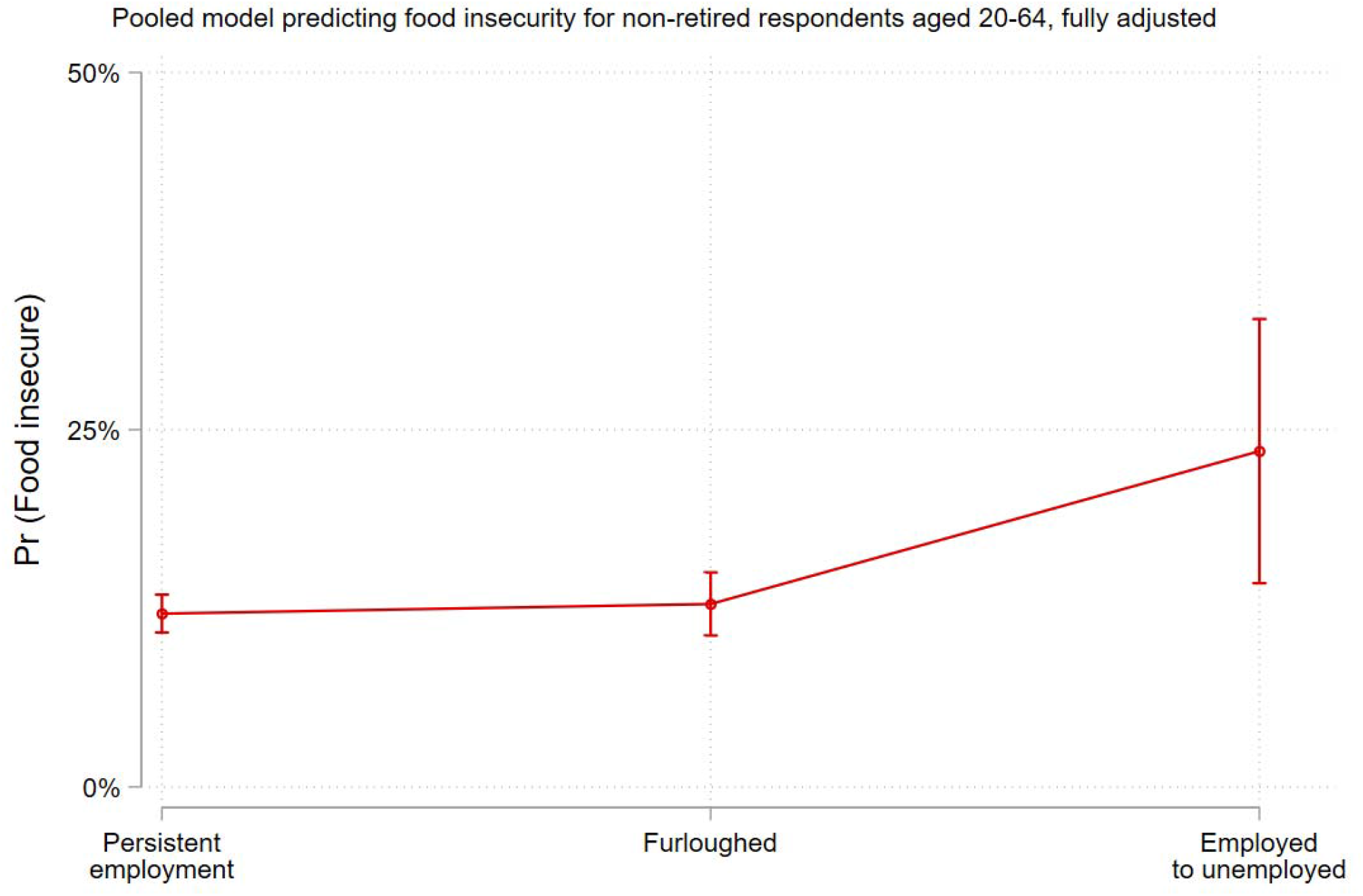
Association between labour force transitions and food insecurity in April and July (N = 5,647), fully adjusted, all respondents employed at baseline (January/February) Note: Estimates are marginal effects derived from a logistic regression model that controls for age, gender, race/ethnicity, number of children at home, highest qualification in 2017-19, equivalized household income in 2015-17, and geographic region. Complex sampling design weights that adjust for non-random attrition are taken into account.

Importantly, furloughed individuals and those benefiting from the equivalent self-employment scheme did not differ significantly in their probability of experiencing food insecurity compared to the persistently employed (OR = 1·07; p = 0·60; 95% CI: 0·83 to 1·37).

## Discussion

These findings document an alarming increase in food insecurity in the United Kingdom during the pandemic. Problems were anticipated^15^ and measures were taken to mitigate them. However, it is clear that they have had limited success. The number of people reporting food insecurity has increased three-fold. All of the groups examined in this analysis have been affected, but some more than others.

Our findings are consistent with a recent report, using different data and food insecurity measures, which found that adults who were working in February 2020 but who reported transitioning to unemployment in May or July were about 2·5 times more likely to report food insecurity compared to those who remained employed (18·5% vs. 7·4%, respectively).^16^ The same study did not find a similar increase for those who had been working in February but who were furloughed in May or June, also suggesting this scheme has mitigated what would have otherwise been a more substantial rise in food insecurity among this group. However, unlike in our study, furloughed respondents reported significantly higher rates of food insecurity relative to those who remained employed. The present study also corroborates findings from a cross-sectional study conducted in early April, again using different data and food insecurity measures, that found higher rates of food insecurity in the UK relative to 2018.^17^ Importantly this latter study relied on items used to measure food insecurity that referred to a 12-month time span in 2018 and then span a 30-day time span in April 2020, a potential source of bias for examining changes in population prevalence over time. Finally, Our study is consistent with recent findings from the USA, finding that food insufficiency among all adults increased three-fold during the COVID pandemic compared to 2019, with African-Americans hardest hit.^18^

As with all analyses of survey data, our study has several limitations. First, our analysis does not provide a causal explanation of the impact of COVID on food insecurity, just an association. Second, the Understanding Society COVID wave was only able to link data to 40% of respondents from wave 8 or 9 of the Understanding Society study. This does not affect internal validity, but complicates assessing representativeness of the UK population. To address this, sampling weights were employed. Further we tested whether those respondents with missing values for food insecurity in both April and July differed from those who did not by all study variables. Table A1 in the appendix presents a logistic regression model predicting missingness on food insecurity in both April and July. Here, we find that the self-employed in July are less likely to be missing relative to the employed, those aged 55-64 are less likely to be missing relative to the 16-24 age group, while those with multiple children are more likely to be missing compared to those with no children. We also detect some regional variations, with those in Scotland, Wales, and the North East of England more likely to be missing compared to those residing in London.

Third, our results do not explicitly identify the disparate factors that might play a role in the associations reported here, such as lockdowns measures intended to mitigate the spread of the virus, or variations in the supply of food, which have been associated with increased food insecurity.^17 19^

Fourth, we only examine the impact of moving from employment to out of employment (furloughed or not), but we do not examine the impact of reducing the number of working hours or reasons for doing so, either voluntarily or not, which might lead to a reduced income and hence to an increased risk of becoming food insecure.

Finally, our analysis is based on self-reported food insecurity of adults included in the survey. This can understate the full magnitude of food insecurity and does not capture the experience of vulnerable groups, such as children and homeless people, for which future research is urgently needed.

Food insecurity can reflect reduced supply, increased price, or reduced spending (either as a consequence of lower income or diversion of spending to other products or to saving, with the latter encouraged by fear of what lies ahead). As noted in the introduction, there were shortages of food in shops immediately after the lockdown in March 2020, largely due to panic buying, but these were rapidly resolved. Food price annual inflation, which had been negative prior to the Brexit referendum but increased rapidly thereafter, has remained at between 1% and 2% between March and July 2020.^20^ The Bank of England’s biannual NMG survey, conducted between 6^th^ April and 1^st^ May, found that while many people experienced reductions in income, even more reduced spending.^21^ Among those employed, 18% experienced a reduction in income but 53% reduced spending. The corresponding figures for those who were self-employed, who were especially severely hit, were 66% and 72%, similar to the figures for those furloughed, at 67% and 66% respectively. Perhaps surprisingly, among the unemployed, many of whom would have continued to receive benefits, the figures were even greater, with 76% both experiencing reduced incomes and reducing spending.

A report for the UK Food Standards Agency offers some qualitative insights into the lived experiences of those affected.^22^ The authors interviewed 20 UK citizens in June 2020, half of whom had been food insecure before the lockdown while the remainder became so after it was implemented. They describe how food insecurity and the need to respond to COVID-19, were superimposed upon many other challenges, including job insecurity, health issues, domestic violence, and debt. Factors contributing to increased risk and vulnerability included an inability to build or draw on financial safety nets, the lack of reliable full-time salaries, working in sectors that did not permit remote working, caring responsibilities that limited alternative sources of income, health, and particularly mental health challenges, and domestic abuse. Restrictions associated with the pandemic contributed to food insecurity in several ways. These included the loss of the inability to join family members for particular meals, such as Sunday lunches, that had previously helped them to stretch their budgets, an inability to afford supermarket delivery fees, reduced access to low-cost shops, competition for low-cost “value” brands that were especially likely to be stockpiled, price increases in shops serving deprived areas, and relying on others to help with their shopping, where a feeling of shame prevented them from asking for the cheapest brands to be purchased. The result was that those interviewed reported relying on food of poor nutritional quality, especially from tins or simple carbohydrates, skipping meals, and compromising on food safety by using out of date products. They also reported emotional problems, linked to the loss of family mealtimes. There was considerable awareness of food banks and food box schemes, but low uptake due to the associated stigma. Statutory welfare provisions, including both pre-existing ones, such as Universal Credit, and those introduced as part of the COVID response were reported as often being difficult to access.

Our results have important implications for policy. First, they demonstrate that the Coronavirus Job Retention scheme appeared to have conferred protection against exposure to food insecurity. However, many employees have been unable to benefit from the scheme, and it is unclear for how long it will be extended. Second, we show that all of the groups we examined have been affected. This is not a problem limited to those on the margins of society. Third, our results are consistent with the hypothesis that, in practical terms, access to affordable food has declined reduced. All age groups, including those of pensionable age whose incomes have largely been unaffected, have experienced increased food insecurity. Steps are needed to provide subsidies or food support, especially since during the pandemic emergency food assistance may not be readily accessible. Taken together our results show that, while COVID is first of all a health crisis, it also has potential to become an escalating social and economic crisis if steps are not taken to protect the weak.

The corresponding author attests that all listed authors meet authorship criteria and that no others meeting the criteria have been omitted.

DS is funded by a Wellcome Trust investigator award and the European Research Council, HRES 313590

## Patient consent for publication

Not applicable

## Data Availability

The data used are publicly available via UK Data Service repository (study numbers 6614 and 8644) and do not require ethical assessment for academic research purposes.

https://www.ukdataservice.ac.uk/

## Conflict of interest

None to declare.

## Appendix

**Table A1.**
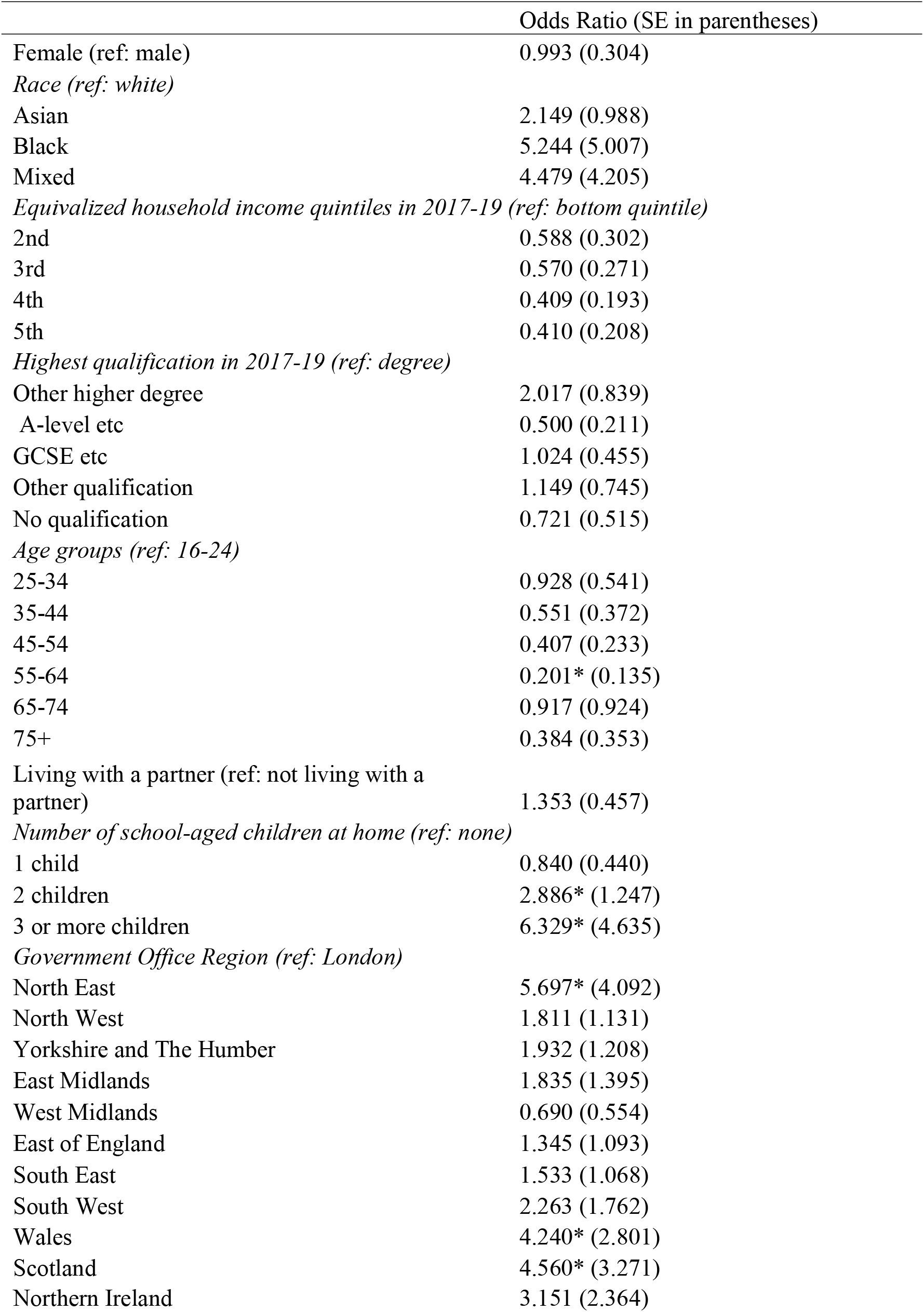
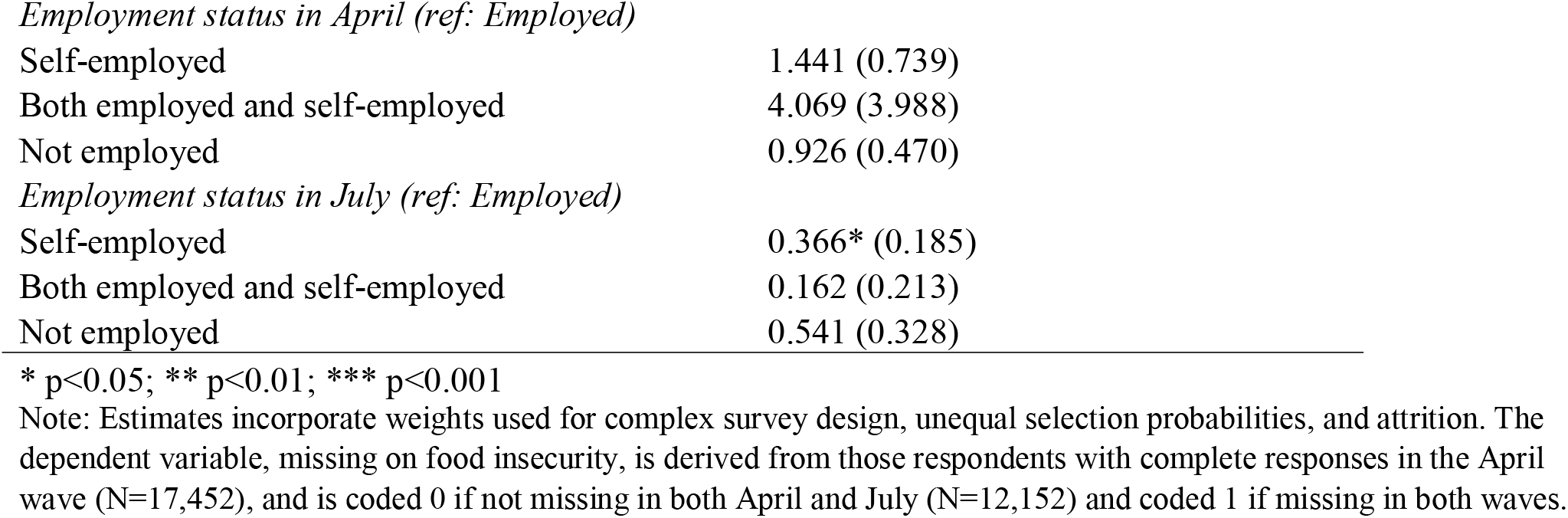
Logistic regression model predicting missingness on food insecurity in both April and July

|  | Odds Ratio (SE in parentheses) |
| --- | --- |
| Female (ref: male) | 0.993 (0.304) |
| *Race (ref: white)* |  |
| Asian | 2.149 (0.988) |
| Black | 5.244 (5.007) |
| Mixed | 4.479 (4.205) |
| *Equivalized household income quintiles in 2017-19 (ref: bottom quintile)* |  |
| 2nd | 0.588 (0.302) |
| 3rd | 0.570 (0.271) |
| 4th | 0.409 (0.193) |
| 5th | 0.410 (0.208) |
| *Highest qualification in 2017-19 (ref: degree)* |  |
| Other higher degree | 2.017 (0.839) |
| A-level etc | 0.500 (0.211) |
| GCSE etc | 1.024 (0.455) |
| Other qualification | 1.149 (0.745) |
| No qualification | 0.721 (0.515) |
| *Age groups (ref: 16-24)* |  |
| 25-34 | 0.928 (0.541) |
| 35-44 | 0.551 (0.372) |
| 45-54 | 0.407 (0.233) |
| 55-64 | 0.201* (0.135) |
| 65-74 | 0.917 (0.924) |
| 75+ | 0.384 (0.353) |
| Living with a partner (ref: not living with a partner) | 1.353 (0.457) |
| *Number of school-aged children at home (ref: none)* |  |
| 1 child | 0.840 (0.440) |
| 2 children | 2.886* (1.247) |
| 3 or more children | 6.329* (4.635) |
| *Government Office Region (ref: London)* |  |
| North East | 5.697* (4.092) |
| North West | 1.811 (1.131) |
| Yorkshire and The Humber | 1.932 (1.208) |
| East Midlands | 1.835 (1.395) |
| West Midlands | 0.690 (0.554) |
| East of England | 1.345 (1.093) |
| South East | 1.533 (1.068) |
| South West | 2.263 (1.762) |
| Wales | 4.240* (2.801) |
| Scotland | 4.560* (3.271) |
| Northern Ireland | 3.151 (2.364) |
| *Employment status in April (ref: Employed)* |  |
| Self-employed | 1.441 (0.739) |
| Both employed and self-employed | 4.069 (3.988) |
| Not employed | 0.926 (0.470) |
| *Employment status in July (ref: Employed)* |  |
| Self-employed | 0.366* (0.185) |
| Both employed and self-employed | 0.162 (0.213) |
| Not employed | 0.541 (0.328) |

\* p<0.05; \*\* p<0.01; \*\*\* p<0.001
Note: Estimates incorporate weights used for complex survey design, unequal selection probabilities, and attrition. The dependent variable, missing on food insecurity, is derived from those respondents with complete responses in the April wave (N=17,452), and is coded 0 if not missing in both April and July (N=12,152) and coded 1 if missing in both waves.

**Figure A1:**
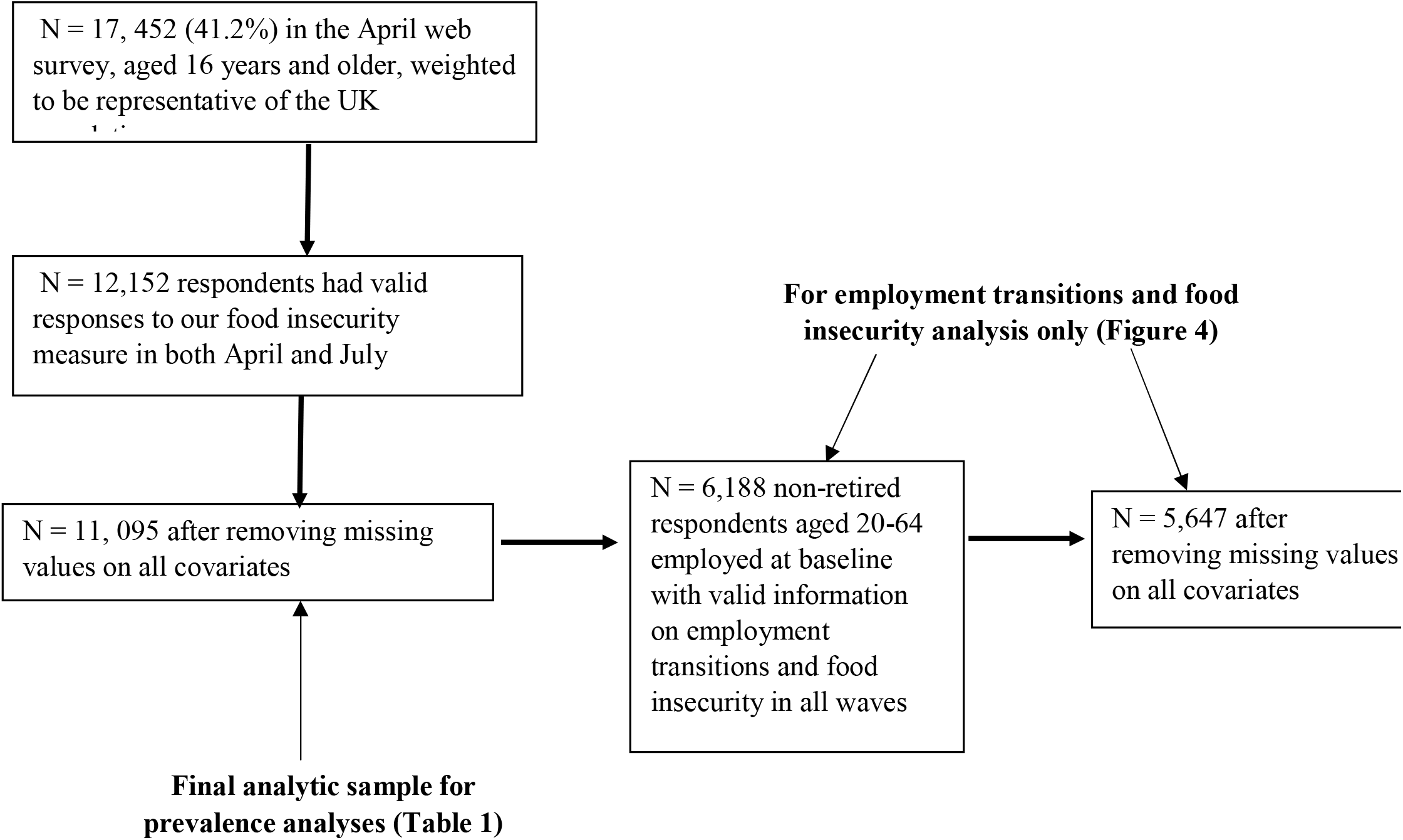
Sample inclusion

## Notes

### Competing Interest Statement

The authors have declared no competing interest.

